# Vietnam National Survey on Parenteral Nutrition Practice in Preterm Neonates: Status, Barriers, and Implications

**DOI:** 10.1101/2024.04.30.24306616

**Authors:** Thu Tinh Nguyen, Phan Minh Nhat Nguyen, Thanh Thien Nguyen, Thi Hieu Vu, Pham Minh Tri Nguyen, Duc Ninh Nguyen

## Abstract

**Background:** Due to high risks of feeding intolerance, preterm infants often receive parenteral nutrition (PN) to ensure sufficient nutrition and energy intake. However, there is a lack of data on the status of clinical PN practice and barriers among neonatal care units in low to middle-income countries like Vietnam. This extensive survey explores the status and barriers of PN practice for preterm infants in neonatal units across Vietnam and identifies the practical implications of enhancing nutritional outcomes in preterm infants.

**Methods:** A multicenter nationwide web-based survey on PN practice in preterm infants was conducted across 114 neonatal units from 61 provinces in Vietnam.

**Results:** Among 114 neonatal units receiving a request for surveys, 104 units (91.2%) from 55 provinces participated. Neonatal units were categorized as level I (2/104, 1.9%), II (39/104, 37.5%), III (56/104, 53.8%), and IV (7/104, 6.8%). We showed that the initiations of PN within the first hour and the first two hours of life occurred in 80.8% (84/104) and 95.2% (99/104) of the units, respectively. The early provision of amino acids (within the first day of life) and lipids (within two days of life) were documented by 85% (89/104) and 82% (84/104) of the respondents, respectively. The initial dose of amino acids ranged from 0.5 to 3 g/kg/day; the dose of amino acids less than 1 g/kg/day was 7.7% (8/104) of respondents; the maximum dose of amino acids ranged from 2 to over 4.5 g/kg/day, with 4 g/kg/day in 47.1% (49/104) of the respondents. The initial dose of lipids was between 0.5 and 2 g/kg/day, frequently 1 g/kg/day, in 51.9% (54/104) of the respondents; the target lipid dose ranged from 3 to 4 g/kg/day in 93.3% (97/104); the maximum target dose for lipid was 4 g/kg/day in 36.5% (38/104) of the respondents. The initial glucose dose was distributed as follows: 46.2% of respondents (48/104) administered 04 mg/kg/minute, 21.2% (22/104) used 05 mg/kg/minute, 28.8% (30/104) used 06 mg/kg/minute, and 3.8% (4/104) used 03 mg/kg/minute. Additionally, 48.1% of respondents (50/104) reported a maximum glucose infusion rate above 13 mg/kg/min and 19.2% (20/104) above 15 mg/kg/min. Nineteen percent (20/104) of the respondents reported lacking micronutrients. Barriers to PN initiation included difficulty in establishing intravenous lines, the absence of standardized protocols, the lack of lipids and micronutrients, infections, and unavailable software supporting neonatologists in calculating nutrition paradigms.

**Conclusions:** This study’s findings highlight the highly variable PN practice across neonatal units in Vietnam. Deviations from current practical guidelines can be explained by various barriers, most of which are modifiable. A monitoring network for nutritional practice status and a database to track the nutritional outcomes of preterm infants in Vietnam are needed.

**Key messages:** *What’s known?:* Recent clinical trials have suggested that parenteral nutrition (PN) is an important factor contributing to both short- and long-term clinical outcomes, including mortality and neurodevelopment, in preterm infants. There are existing well-developed PN guidelines for preterm infants in high-income countries.

*What’s new?:* PN practice varies across neonatal units in Vietnam, particularly in amino acids, lipids, and micronutrient provision. Deviations from the current practical guidelines are related to various barriers. Common barriers include difficulty in establishing intravenous lines, the absence of standardized protocols, the unavailability of lipids and particularly micronutrients, and the lack of software supporting neonatologists in calculating nutritional paradigms. Fortunately, most of these barriers are modifiable. Establishing a monitoring network for nutritional practice status and developing a database to track the nutritional outcomes of preterm infants in Vietnam is necessary.

## INTRODUCTION

Approximately 4-16% of total live births are preterm births (< 37 weeks of gestation) [1], which leads to multiple serious complications with high mortality and morbidity during the neonatal period. These complications seem to be universal in both high-income and low-to-middle-income countries and contribute to around one-third of neonatal deaths[2]. Complications related to preterm birth also cause long-term health consequences, such as an increased risk of hypertension, insulin resistance, and neurodevelopmental disabilities later in life [3,4]. As the survival rate of preterm infants has increased in low-to-middle-income countries like Vietnam, preterm birth likely imposes a greater burden and impact on society[4].

Recent clinical trials have shown that parenteral nutrition (PN) is important for clinical and neurodevelopmental outcomes in preterm infants. For instance, delayed or insufficient nutrient provision may lead to energy deficiency, elevating the risk of complications such as extrauterine growth retardation, cortical myelination deficiency, and neurodevelopmental impairment[3,5,6]. Ensuring timely and sufficient nutrition is key to enhancing survival rate and quality of life for premature infants[5,7].

Despite the availability of PN guidelines for preterm infants in high-income countries[8-10], it is unclear how clinical practice is implemented at neonatal care units in Vietnam. Reports from NICUs worldwide have shown frequent inadequate nutritional intake in early life, and the reasons behind this are multifactorial [11-13]. These may include variable PN practices among units, but it is possible that preterm infants in low-to-middle-income countries need a distinct PN guideline from the one applied in high-income countries[4].

From this background, we conducted a survey to assess the status and barriers to PN practice in Vietnamese NICUs. The findings aim to help develop solutions for enhancing PN practices for preterm infants in low-to-middle-income like Vietnam.

## METHODS

The study received ethical approval from the Institutional Review Board of Children’s Hospital 2 on May 15th, 2023. Project No.: 211/GCN-BVNĐ2.

This cross-sectional survey was designed to capture insights into current practice status and barriers associated with PN among neonatologists. A survey questionnaire, developed using Microsoft Forms, included twenty-nine multiple-choice questions and four optional free-text items, enabling comprehensive data gathering on the topic. In addition, the responses were anonymous and did not include patient data.

Prior to the main survey, the questionnaire was pretested with pediatric postgraduate students to ensure clarity, detail, and a logical sequence of questions. This pilot survey provided valuable feedback that informed the refinement of the survey items. The list of target respondents included neonatal units from 61 provinces in Vietnam known to administer PN to preterm neonates. Two of the 63 provinces were excluded due to a lack of parenteral nutrition services.

Pre-ordered PN is unavailable in Vietnam. Instead, PN is mixed either in the pharmacy or directly within units, depending on local policy and available facilities. Neonatologists adjust the orders for PN for each infant daily, typically during morning or afternoon rounds.

In total, 114 anonymous online questionnaires were distributed through email, messages, and social messaging platforms to NICUs across these 61 provinces over one month from June 1st to June 30th, 2021. The collection of responses was collected through Microsoft Forms. The initial communication for the survey detailed the elements of written consent, and the act of response was taken as consent to participate. This introductory message also advised that a representative from each unit, preferably a senior neonatologist, should complete the survey to avoid duplicate responses and more reliable data. Throughout the survey period, weekly reminders were sent by the data management team to maximize the response rate. The average time required to complete the survey, as pretested during the pilot with postgraduate students, was approximately five minutes.

All data were anonymized to protect the identities of the participating neonatologists. The classification of NICUs was based on the American Academy of Pediatrics (AAP) level of neonatal care [14]. Level I (Essential care): Stabilize and provide care for newborns ≥ 35 weeks of gestation; stabilize newborns < 35 weeks. Level II (specialty care): Level I + provides care for newborns≥ 32 weeks of gestation and weighing ≥ 1500g and provides mechanical ventilation for a brief duration (<24h) or continuous positive airway pressure. Level III (Intensive care): Level II + provides care for infants born at all gestational ages, critical care (advanced respiratory-cardiovascular support: mechanical ventilation, surfactant, inhaled nitric oxide (iNO)), and surgical. Level IV (Regional Neonatal intensive care unit): Level III + Provide surgical repair of complex congenital or acquired conditions ((e.g., complex congenital cardiac malformations), provide outreach education).

For statistical analysis, data were first exported to Excel and subsequently imported into R statistical software (Rstudio 2023.12.1+402 for MacOS). The barriers in PN were collected from free text responses, which had to be read through and coded into variations. Descriptive statistics were utilized to summarize the survey findings, and the qualitative data were presented in terms of both the frequency and percentage of responses, as well as the identification of barriers encountered in PN practice.

## RESULTS

Representatives from 104 NICUs across 55 provinces in Vietnam answered the survey. The response rate was 104 responses out of 114 distributed surveys (91.2%).

The participating NICUs were categorized based on levels of neonatal care: Level I, 2 out of 104 (1.9%); Level II, 39 out of 104 (37.5%); Level III, 56 out of 104 (53.8%), and Level IV, 7 out of 104 (6.8%) (see Table 1)

**Table 1:**
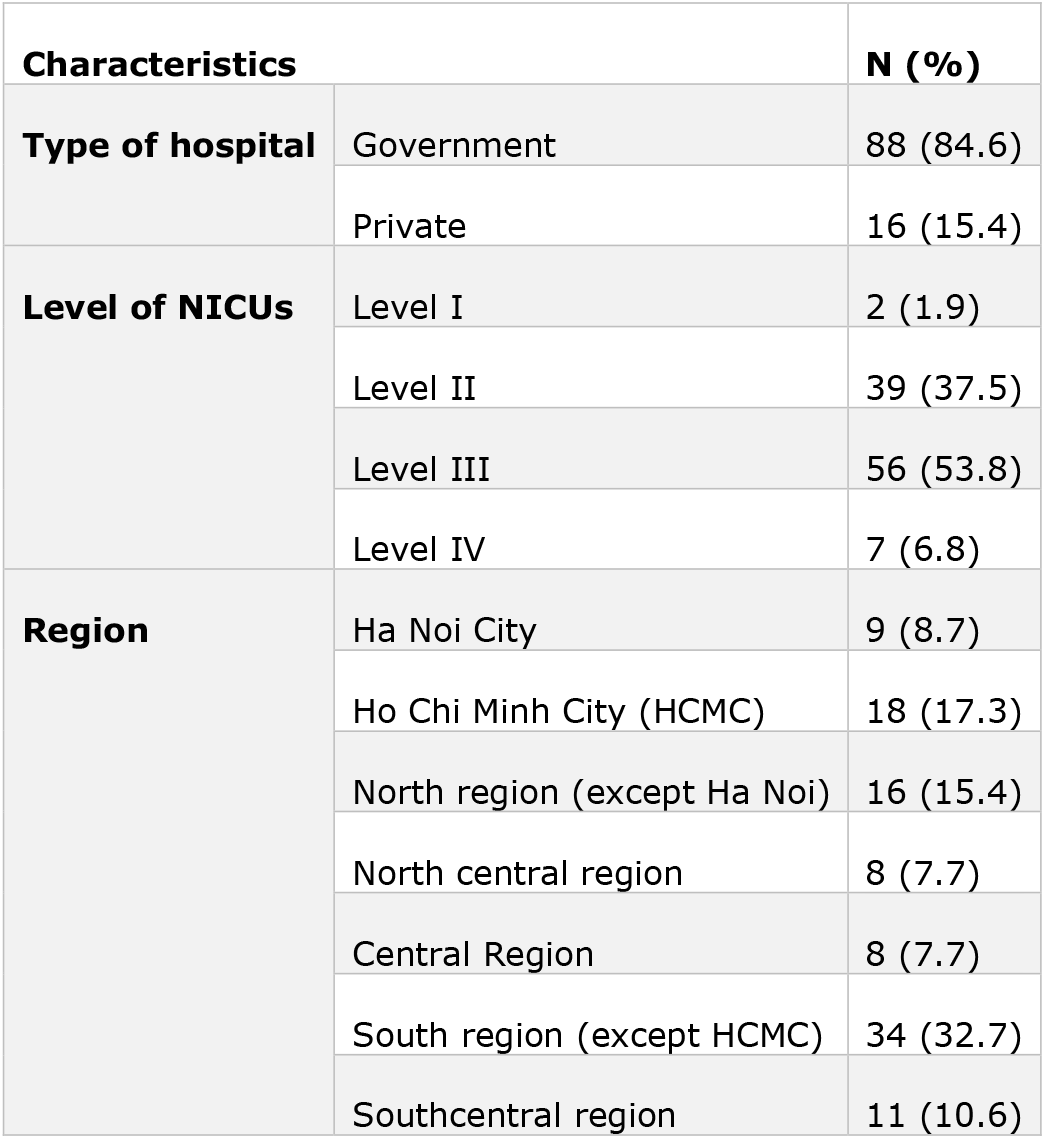
Characteristics of neonatal intensive care units responding to a survey on parenteral nutrition (N = 104). Data are presented as N (%).

| **Characteristics** |  | **N (%)** |
| --- | --- | --- |
| **Type of hospital** | Government | 88 (84.6) |
|  | Private | 16 (15.4) |
| **Level of NICUs** | Level I | 2 (1.9) |
|  | Level II | 39 (37.5) |
|  | Level III | 56 (53.8) |
|  | Level IV | 7 (6.8) |
| **Region** | Ha Noi City | 9 (8.7) |
|  | Ho Chi Minh City (HCMC) | 18 (17.3) |
|  | North region (except Ha Noi) | 16 (15.4) |
|  | North central region | 8 (7.7) |
|  | Central Region | 8 (7.7) |
|  | South region (except HCMC) | 34 (32.7) |
|  | Southcentral region | 11 (10.6) |

The initiations of PN for preterm infants within the first hour of life and the first two hours of life were 80.8% (84 out of 104) and 95.2% (99 out of 104), respectively.

### Amino acids

Over 85 percent (89 out of 104) of the respondents provided parenteral amino acids within the first day of life, while 31.7% (33 out of 104) and 4.8% (5 out of 104) initiated on the second day and third day of life, respectively. The initial dose of amino acids ranged from 0.5 to 3 g/kg/day. The dose of amino acids less than 1 g/kg/day was reported in 7.7% (8 out of 104) of the respondents. The maximum dose of amino acids ranged from 2 to 4.5 g/kg/day, with 4 g/kg/day reported in 49 of 104 (47.1%) of the respondents (Table 2).

**Table 2:** Current practice of parenteral nutrition (amino acids and lipids) in neonatal intensive care units responding to the surveys (N = 104). Data are presented as N (%).

|  |  | **Number of units (percent)** |  |
| --- | --- | --- | --- |
| **Variables** |  | **Amino acids** | **Lipids** |
| **Initial time (hours), n (%)** | within 24 | 89 (85.6) | 59 (56.7) |
|  | 24 to 48 | 11 (10.6) | 26 (25.0) |
|  | Above 48 | 04 (3.8) | 19 (18.3) |
| **Initial dose (g/kg/day), n (%)** | 0.5 | 08 (7.7) | 36 (34.6) |
|  | 1.0 | 26 (25.0) | 54 (51.9) |
|  | 1.5 | 35 (33.7) | 07 (6.7) |
|  | 2.0 | 28 (26.9) | 06 (5.8) |
|  | 2.5 | 2 (1.9) | - |
|  | 3.0 | 5 (4.8) | 1 (1.0) |
| **Maximum dose (g/kg/day)** | 2.0 | 1 (1.0) | 3 (2.9) |
|  | 2.5 | 03 (2.9) | 3 (2.9) |
|  | 3.0 | 08 (7.7) | 37 (35.6) |
|  | 3.5 | 28 (26.9) | 22 (21.2) |
|  | 4.0 | 49 (47.1) | 38 (36.5) |
|  | 4.5 | 12 (11.5) | 1 (1.0) |
|  | > 4.5 | 3 (2.9) | - |

### Lipids

Eighty-two percent (85 out of 104) of the respondents reported the initiation of parenteral lipids within two days of life; of those, 69.4% (59 out of 85) initiated within the first 24 hours of life. The initial dose was between 0.5 and 2.0 g/kg/day, frequently 1 g/kg/day, except in 1 respondent in which the dose was 3 g/kg/day, in 51.9% (54 out of 104). One respondent (1%) reported that lipids were not available in the NICU. The target lipids dose of 3 to 4 g/kg/day was reported by 93.3% (97 out of 104) of respondents. Additionally, the maximum target dose for lipids was 4 g/kg/d for 36.5% (38 out of 104) of respondents (Table 2)

### Glucose

The initial glucose dose was distributed as follows: 46.2% (48 out of 104) of the respondents used 4 mg/kg/minute, 21.2% (22 out of 104) used 5 mg/kg/minute, 28.8% (30 out of 104) used 6 mg/kg/minute, and 3.8% (4 out of 104) used 3 mg/kg/minute. Additionally, the maximum rate of glucose infusion above 13 mg/kg/min and 15 mg/kg/min were reported by 50 (48.1%) and 20 (19.2%) respondents, respectively.

Nineteen percent (20 out of 104) of the respondents reported lacking parenteral micronutrients.

### Barriers

Figure 1 Details the barriers that have deterred the respondents from applying their knowledge to current clinical practice. Among these, difficulties in establishing vascular access, the absence of local PN guidelines and lack of micronutrients are the most common reported barriers.

**Figure 1:**
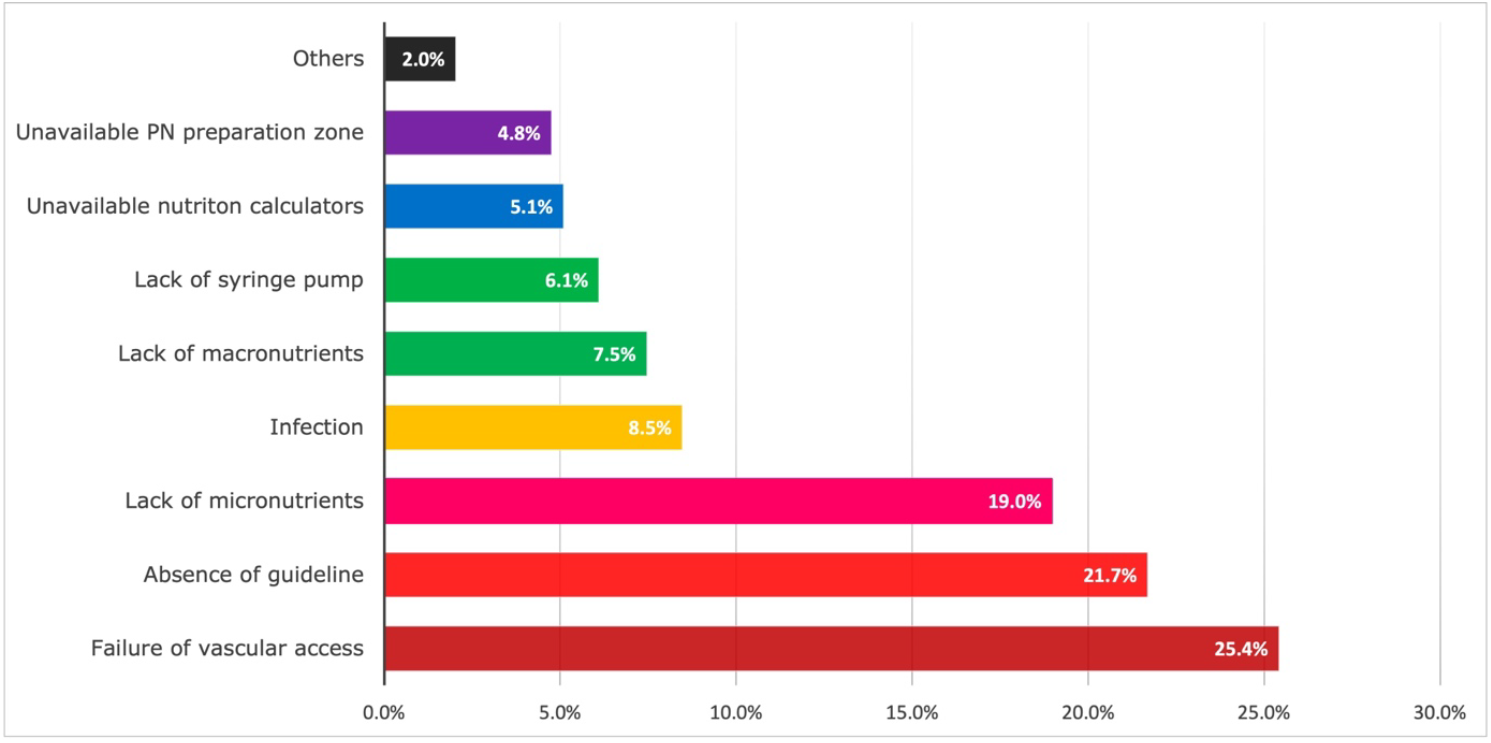
Main barriers affecting parenteral nutrition practice among NICUs in Vietnam. A total of 295 opinions received from 104 respondents were categorized into eight types of barriers. The percentages were calculated based on a total of opinions (N = 295).

A total of 295 opinions received from 104 respondents were categorized into eight types of barriers (Figure 1)

## DISCUSSION

It is well known that PN is associated with multiple clinical and neurodevelopmental outcomes in preterm infants [3,5,6,15]. The PN practices are highly variable among neonatal units, and units in low-to-middle-income countries may need distinct PN guidelines[4]. Here, we conducted a national survey of PN practice across 55 provinces in Vietnam with 91.3% (95 out of 104) of NICUs from levels of care II to III, which are in the general hospitals. Many of these NICUs lack sufficient facilities and staff that care for preterm infants. The most recent European and U.S. guidelines recommended that PN should be initiated early, within the first 1 to 2 hours of life in preterm infants [9,10,16,17], and our survey results reported a high level of compliance.

The survey documented that 85.6% (89 out of 104) of NICUs provided amino acids (AA) within 24 hours of life (on the PN was initiated), consistent with existing guidelines[9,10,16-18]. Still, the remaining NICUs (14.4%) deferred providing AA. Reasons for this include the need for medical consultation and concerns about kidney function of preterm infants. According to a study in premature infants <30 weeks of gestation, levels of creatinine remained in the normal range following PN with recommended protein intake during the first three weeks of life [19]. In principle, the provision of AA aims to mimic the AA accretion by the fetus. The initial AA dose recommended in clinical trials and guidelines was 1-2 g/kg/day, which is expected to prevent a net negative nitrogen balance [9,18,20]. Our survey revealed that the initial dose of AA ranged from 1 to 2 g/kg/day in 85.6% (89 out of 104) NICUs. For the maximum dose of AA, this varies in previous clinical trials from 2.5 to 4.5 g/kg/day [17]. Current evidence also shows limited difference in clinical outcomes between a maximum AA dose of 3.5 vs 4 g/kg/day [5,17]. Our survey showed that the maximum dose of AA is consistent with the recommendations (ranging from 3.5 to 4 g/kg/day) in 74% (77 out of 104) of the NICUs.

The provision of parenteral lipids aims to improve growth and prevent an essential fatty acid deficiency (EFAD). The survey documented the use of parenteral lipids in 99% (103 out of 104) of the NICUs. Lipids were initiated early on the first 2 days of life for 81.7% (85 out of 104) of the respondents. The remaining 18% (19 out of 104) of the NICUs started lipids on three days of life because they believed that lipids need to be administered separately from PN. In fact, lipids can be infused along with parenteral nutrition via 3-way stopcocks. The recommended initial dose of lipids ranges from 0.5 to 2 g/kg/day, whereas our study showed that 86.5% (90 out of 104) of the units used 0.5-1 g/kg/day. The maximal target dose of lipids was 4 g/kg/day for a minority of respondents, at 35.6% (37 out of 104). This minority could be explained by the low rate of extremely low birth weight infants in their NICUs. The specific composition of lipids might play a role of benefit in enhancing growth outcomes. The ideal lipid emulsion should contain n-6 and n-3 fatty acids (fish oil), antioxidant agents (e.g., tocopherol and monounsaturated fatty acids), and medium-chain fatty acids [9,16]. Clinically, most units use 20% lipid emulsions (instead of 10%) because it provides more energy than 10% emulsions (2 kcal/ml vs. 1,1 kcal/ml), contains a lower phospholipid content, with limited increase in plasma triglycerides [10].

The survey documented the maximal target dose of glucose over 13 mg/kg/min in 48.1% (50 out of 104). The glucose infusion > 12 mg/kg/min may overwhelm the maximal capacity of glucose oxidation of the liver in preterm infants. In contrast, the rate of 4 mg/kg/min is the estimated rate of glucose use by the brain [8]. In addition, a normal daily breastmilk intake of about 150 ml/kg/day can provide a glucose rate of 4 mg/kg/min, and this was the threshold for initial glucose infusion documented by 46.2% (48 out of 104) of NICUs. Despite the demand for high energy for growth in very low-birth-weight infants, they do not often tolerate the recommended glucose dose during the first few days of life. These infants have high risks of hyperglycemia, explaining the balance of using an initial glucose infusion of 4 mg/kg/min. On the other hand, other groups of preterm infants are recommended to start with a glucose regimen of 5-6 mg/kg /min [8].

There were multiple recorded barriers affecting PN practice among NICUs in Vietnam. Most of the mentioned barriers include failure of venous access, lack of parenteral protocols or consensus guidelines, unavailable methods of PN calculation, and unavailable micronutrient products. Some barriers can actually be resolved in the near future by developing national PN guidelines, improving PICC insertion procedures, and developing computer software to support decision-making [21]. Simultaneously, there is an urgent need for the establishment of a collaborative nutritional network and a national database to evaluate nutritional outcomes in the future.

## CONCLUSIONS

The findings of this study highlight great variation in PN practice in Vietnam, particularly in amino acids and micronutrient provision. Deviations from current practical guidelines can be explained by various barriers, most of which are modifiable. There is an urgent need to establish a monitoring network for nutritional practice status and develop a database to track the nutritional outcomes of preterm infants in Vietnam. Future surveys will be conducted to determine the effects of interventions on barriers and nutritional outcomes of preterm infants in Vietnam.

## Data Availability

The data presented in this study are available on reasonable request from the corresponding author.

## Acknowledgments

We would like to acknowledge all our colleagues who enrolled in the study. We want to acknowledge our supporting colleagues at the University of Medicine and Pharmacy at Ho Chi Minh City and Children’s Hospital 2 in Ho Chi Minh City, Vietnam.

## Author contributions

TTN, PMNN, TTN, THV, PMTN developed the questionnaires. All authors disseminated the questionnaire via established neonatal networks and online messaging platforms. All authors performed statistical analyses and drafted the manuscript. Supervision: TTN. All authors contributed to data interpretation, manuscript revision, and approval of the final manuscript version.

## Data Availability Statement

The data presented in this study are available on reasonable request from the corresponding author.

